# Evaluating quality improvement at scale: routine reporting for executive board governance in a UK National Health Service organisation

**DOI:** 10.1101/2020.02.13.20022475

**Authors:** Kia-Chong Chua, Claire Henderson, Barbara Grey, Michael Holland, Nick Sevdalis

## Abstract

**Purpose:** Quality improvement (QI) in healthcare is a cultural transformation process that requires long-term commitment from the executive board. As such, an overview of QI applications and their impact needs to be made routinely visible. We explored how routine reporting could be developed for QI governance.

**Design:** We developed a retrospective evaluation of QI projects in an NHS healthcare organisation. The evaluation was conducted as an online survey so that the data accrual process resembled routine reporting to help identify implementation challenges. A purposive sample of QI projects was identified to maximise contrast between projects that were or were not successful as determined by the resident QI team. To hone strategic focus in what should be reported, we also compared factors that might affect project outcomes.

**Findings:** Out of 52 QI projects, 10 led to a change in routine practice (‘adoption’). Details of project outcomes were limited. Project team outcomes, indicative of capacity building, were not systematically documented. Service user involvement, quality of measurement plan, fidelity of plan-do-study-act (PDSA) cycles had a major impact on adoption. We discussed how routine visibility of these factors may aid QI governance.

**Originality:** Designing a routine reporting framework is an iterative process involving continual dialogue with frontline staff and improvement specialists to navigate data accrual demands. We demonstrated how a retrospective evaluation, as in this study, can yield empirical insights to support dialogue around QI governance, thereby honing the implementation science of QI in a healthcare organisation.

## BACKGROUND

A growing number of health care provider organisations in the UK National Health Service (NHS) are adopting quality improvement (QI) strategies across their organisations (Ross and Naylor, 2017). The Care Quality Commission (CQC), an independent regulator of health and social care services in England, noted a trend after recent inspections that NHS provider organisations with outstanding CQC ratings have applied QI at scale (Care Quality Commission, 2018b, Care Quality Commission, 2018a). The systematic application of QI at scale in NHS provider organisations has required concerted investment to sustain a process of cultural transformation. To monitor the developmental stages of this process, an organisation-wide picture of QI applications and their impact is needed. This helps the organisation’s executive board commit to a long-term perspective on investments made in QI infrastructure, a critical theme in emerging accounts of QI success in NHS provider organisations (Ross and Naylor, 2017, Shah and Course, 2018).

With prevailing pressures on NHS resources (Dunn et al., 2016, Smithson et al., 2015, Gilburt, 2015), exacerbated by the COVID-19 pandemic, sustaining long-term commitment of the executive board would require routine reports to increase visibility of the QI success and progress in cultural transformation. Amidst growing literature on hospital board governance for QI (Jones et al., 2017), not much is known about what routine reporting should offer to be useful for this governance. Firstly, routine reporting needs systematic design that addresses staff capacity building, particularly in developmental stages of embedding a culture of continuous improvement within an organisation. The level of technical skills needed for measurement, data collection and analysis is commonly underestimated in QI, and generally not sufficient in frontline NHS staff despite some training provision (Woodcock et al., 2019). Moreover, routinely collected data are often not as clean or well set up as originally anticipated, often requiring extensive effort to bring them up to a standard suitable for use (Woodcock et al., 2019). This means that an organisation-wide picture of QI applications and their impact cannot be readily constructed by aggregating project reports of impact or success. Even for case studies with proper design and adequate analysis, they would not offer an organisation-wide picture of QI success and cultural transformation.

There is, therefore, a need for constructing a routine reporting framework that can help inform the executive board and sustain long-term commitment in utilising QI methodology to achieve cultural transformation within a healthcare organisation. This study aims to start addressing this gap. We report a pilot evaluation of QI activity within a large mental health NHS Trust to develop the content of routine reporting. To hone strategic focus in what should be reported, we also compared factors that might affect project outcomes. We review evaluation findings, alongside data quality, to inform on design considerations and implementation challenges of routine reporting for the Trust’s executive board.

## METHOD

### Setting

An NHS organisation that provides specialist care for mental health in South London established a resident QI team in 2016 with the mandate to foster a continuous improvement culture. The team supports QI projects led by frontline staff through training and coaching on QI methodology (e.g., Model for Improvement, driver diagrams, measurement plans, Plan-Do-Study-Act (PDSA) cycles). In partnership with an academic institution, the NHS provider organisation also adopted a researcher-in-residence model that embeds an academic faculty in the resident QI team to support data and evaluation needs. With a steady rise of QI activities within the organisation between 2016 and 2018, the executive board requested an evaluation of these nascent developments.

### Study Design

To explore what should be included for an executive board report on organisation-wide QI programme, a scoping exercise was first initiated by the researcher-in-residence to develop a proposal of evaluation content. This involved individual consultations with three improvement specialist colleagues in the resident QI team, coupled with a rapid evidence scan of peer-reviewed and grey literature on what constitutes success for QI in healthcare. Key concerns were raised with time resources, staff turnover, service user involvement, and outcomes like whether project ideas were adopted and spread. The literature also drew attention to the fidelity of PDSA applications and implementation of data or measurement plan.

Following this process, a list of items was generated and circulated among the entire team of six improvement specialists in the resident QI team for formative feedback on content relevance, acceptability and feasibility issues. This feedback was combined with inputs from two academic colleagues with expertise in implementation and improvement sciences. A revised version was then piloted with two colleagues in the resident QI team. Based on the finalised content, an online template was constructed (online supplement) so that the data accrual process simulated what would be required for routine reporting to the executive. This was intended to surface challenges in routine reporting and guide design considerations.

With approximately 200 QI projects initiated between 2016-18, retrospective information retrieval was undertaken with a purposive sample for feasibility reasons. Each improvement specialist of the Trust’s resident QI team was requested to complete the online template for up to five QI projects that they considered as ‘successful’ and up to another five that they considered as ‘unsuccessful’. Specifically, they were asked to rely on their own assessment of what did and did not work to maximise the gradient of contrast across the selection of QI projects. We decided on this approach because there is no established definition of ‘success’ in QI (Morganti et al., 2012). Instead of imposing a ‘work-as-imagined’ criterion in the absence of consensus, we thus aim to gain insights on staff perceptions of success in terms of ‘work-as-done’ (Hollnagel, 2017).

### Measures

The online template aimed to serve a dual purpose of accountability and feedback. Accountability measures focus on project outcomes, costs and benefits. Feedback measures focus on contextual, input, and process factors that may affect project outcomes. To offer an overview of a diverse range of QI projects, we operationalised the measures at a conceptual level that allow comparisons between projects despite heterogeneous and highly localised QI project aims.

In terms of project outcomes, we studied whether projects: (1) reached formal completion; (2) achieved their aims; (3) led to a change in routine practice (adoption); and (4) triggered similar projects beyond the site of their original conception (spread). To form a picture of cost and benefit amidst considerable variation in documentation, we probed on whether it was possible to quantify improvement in terms of cost savings and whether this had been attempted. As a practical measure of cost, we measured the amount of contact with the resident QI team in terms of meetings, site visits, and email correspondence. In terms of benefit, we sought an estimate of the number of staff and service users who benefitted directly from the change introduced by the QI project. We also probed on two types of project milestones: (1) the extent and form in which project teams disseminated their findings and (2) whether project team members went on to develop new QI projects. These latter measures help provide an account of capacity building across projects. As an organisational management philosophy, QI recognises the critical need to empower frontline staff to learn and participate in continuous improvement in face of escalating complexity and change (Blumenthal and Kilo, 1998). As such, capacity building is a pertinent milestone in efforts to move beyond performance assurance and cultivate an organisational culture of learning.

To enable feedback on what may facilitate or impede QI project success, we explored contextual, input and process factors. Contextual factors refer to organisational conditions that are not within the influence of QI project teams yet can impact on project outcomes and success. Besides setting (inpatient / community care), the resident QI team drew particular attention to contextual aspects such as whether a project was the team’s first ever QI project undertaken, and whether protected time for QI activities was officially sanctioned. Input factors comprised team characteristics and staff turnover. Process factors referred to actions or decisions of the project teams. They comprised stakeholder engagement, PDSA cycles and measurement plans implemented.

### Analysis

We first enumerated project outcomes in terms of whether they reached formal completion, achieved their aims, introduced change ideas that were adopted in routine practice (adoption), and triggered similar projects at other sites (spread). Thereafter, we focused on adoption as the dependent variable in univariable logistic regression models that compared the odds of adoption for each contextual, input, and process factors. An odds ratio (OR) smaller than 1.5 was considered to be a small effect size, whereas OR > 5.0 was considered to be a large effect size (Chen et al., 2010).

## RESULTS

The resident team of six improvement specialists reported 52 QI projects across five boroughs of London (UK) served by the NHS provider organisation (Table 1). Thirty projects were conceived by community mental health teams and the remaining (n=22) by inpatient care teams. Of the three Quality Priority themes (South London and Maudsley NHS Foundation Trust, 2017: patient safety, clinical effectiveness, and patient experience) improving clinical effectiveness was the most common focus in project aims (29 / 52). A minority of projects focused on multiple priorities at once (6 community mental health and 7 inpatient care projects).

**Table 1.**
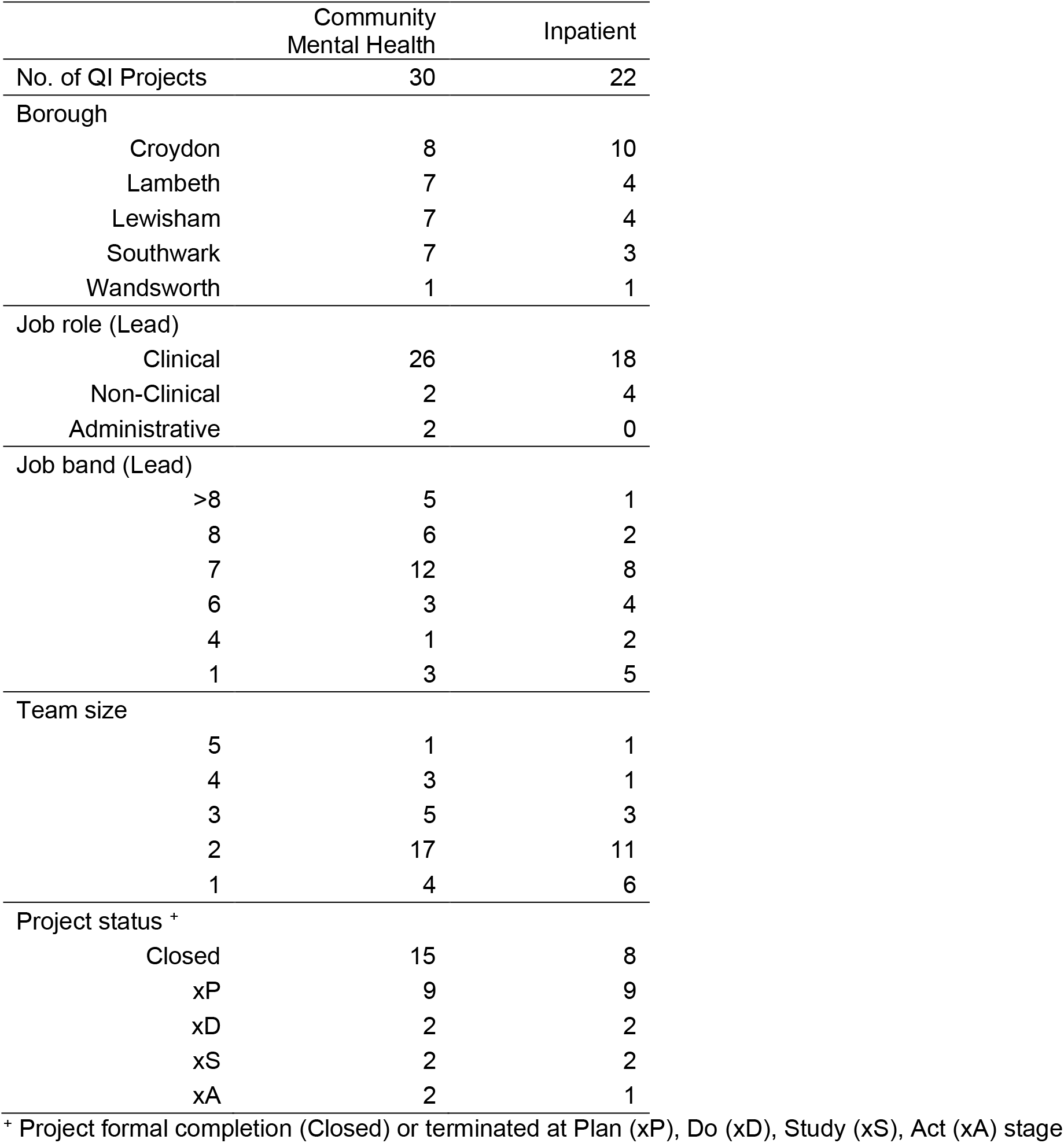
Study sample (n=52) of quality improvement projects in South London and Maudsley NHS Foundation Trust (2016 – 2018)

Before making recommendations (in Discussion) on which accountability and feedback measures could be reported to the executive board, we briefly highlight in this section how each result may offer potentially actionable insights for QI governance.

As a result of purposive sampling, approximately half of the 52 projects were reported by the QI resident team to have reached formal completion (Table 2). In the remaining half of the projects that were terminated prematurely, most were at the planning stage. Among 23 projects that reached formal completion, 12 achieved their aim, but 13 were reported to have led to a change in routine practice (adoption). We observed a similar inconsistency among projects that were terminated prematurely. A plausible explanation could be that some projects were organisation-wide directives that were adopted or spread across services despite not completing or achieving their aims at specific sites. This reflects a gap between “work-as-imagined” and “work-as-done”(Hollnagel, 2017). In terms of QI governance, such a gap may be a useful reference point to mark the stage of cultural transformation amidst growing applications of QI within the organisation.

**Table 2.**
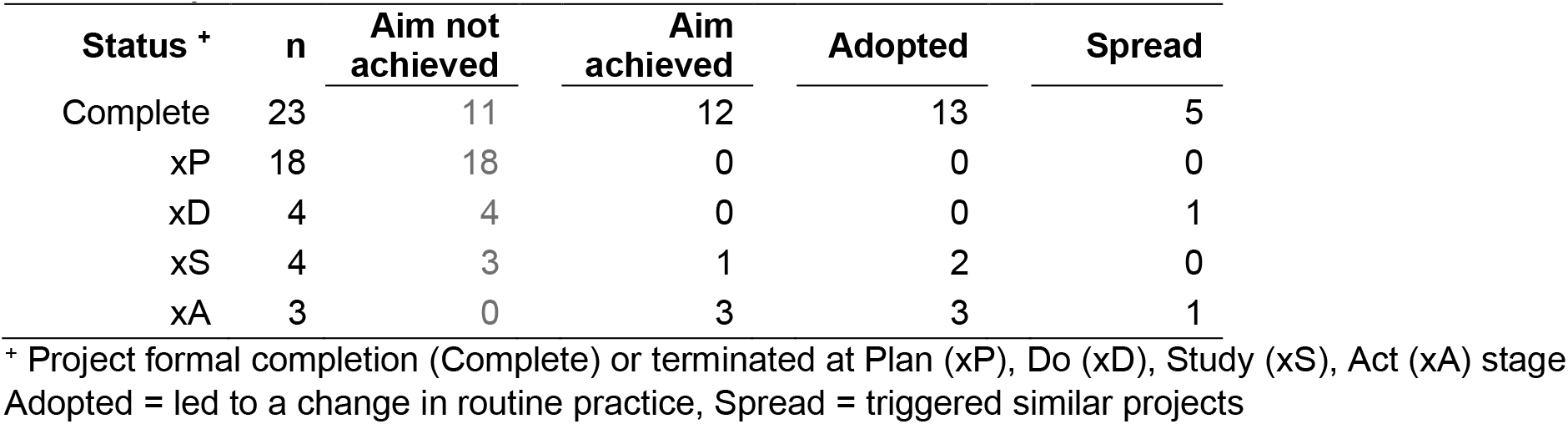
Project outcomes

| Status + | n | Aim not achieved | Aim achieved | Adopted | Spread |
| --- | --- | --- | --- | --- | --- |
| Complete | 23 | 11 | 12 | 13 | 5 |
| xP | 18 | 18 | 0 | 0 | 0 |
| xD | 4 | 4 | 0 | 0 | 1 |
| xS | 4 | 3 | 1 | 2 | 0 |
| xA | 3 | 0 | 3 | 3 | 1 |

+ Project formal completion (Complete) or terminated at Plan (xP), Do (xD), Study (xS), Act (xA) stage Adopted = led to a change in routine practice, Spread = triggered similar projects

Table 3 shows project duration and amount of contact with the QI resident team. Projects typically lasted almost 6 months whether they reached formal completion (median = 5.8 months) or terminated prematurely (median = 5.1 – 6.0 months). The only exceptions were those that terminated at the planning stage (median = 1.8 months). Similarly, meetings and site visits typically took place monthly regardless of project trajectories. The resource burden incurred by projects that terminated prematurely is significant given their numbers. In terms of QI governance, formulating gatekeeping policy directives may help ensure that competing demands for the QI resident team do not wear down available resources.

**Table 3.** Project duration and amount of contact with QI resident team

| Status + | n | Duration (Months) |  |  | Meetings |  |  | Site Visits |  |  | Emails |  |  |
| --- | --- | --- | --- | --- | --- | --- | --- | --- | --- | --- | --- | --- | --- |
|  |  | Md | M | SD | Md | M | SD | Md | M | SD | Md | M | SD |
| Closed | 23 | 5.8 | 5.1 | 3.6 | 5.0 | 5.2 | 3.3 | 4.0 | 5.5 | 5.3 | 25.0 | 25.3 | 20.6 |
| xP | 18 | 1.8 | 2.1 | 1.6 | 2.0 | 1.5 | 1.2 | 1.0 | 1.6 | 1.9 | 7.5 | 8.2 | 4.6 |
| xD | 4 | 5.1 | 5.0 | 3.1 | 3.5 | 3.5 | 1.3 | 4.5 | 5.0 | 4.7 | 20.0 | 22.8 | 16.1 |
| xS | 4 | 5.8 | 5.7 | 1.5 | 4.5 | 4.5 | 1.3 | 4.0 | 4.3 | 3.0 | 20.5 | 29.8 | 27.8 |
| xA | 3 | 6.0 | 5.8 | 1.2 | 4.0 | 4.7 | 3.1 | 4.0 | 4.7 | 3.1 | 20.0 | 28.3 | 28.4 |

+ Project formal completion (Closed) or terminated at Plan (xP), Do (xD), Study (xS), Act (xA) stage Md: Median, M: Mean, SD: Standard Deviation

In contrast to time use data, retrospective estimates offered sparse data across various indicators of project benefit. Approximately two-thirds of the projects did not have estimates of the number of service users and staff who directly benefitted from the change introduced. For a similar proportion, it was not possible to quantify improvement in terms of cost savings. In the remaining one third, it was possible but not attempted. It was not known for three quarters of the projects whether they disseminated their results. Only a small number of projects disseminated their results in a form of newsletter publication locally or beyond (n= 4 and 6, respectively). Team members of 8 projects were known to have developed 12 new projects. This was unclear for the remaining 44 project teams. Limited visibility of the growth and impact of QI in the organisation may erode long term commitment of the executive board. In terms of QI governance, our retrospective data at large points to a need for systematic monitoring to offer routine visibility.

Table 4 shows how the odds of adoption were influenced by each contextual, input, and process factors. Of the 52 QI projects led by frontline NHS staff, 13 achieved this goal. However, in light of the gap between “work as imagined” and “work as done”, we used a more stringent definition of adoption by labelling projects as successful (n=10) only if they met all three conditions: (a) formal project completion; (b) achieved project aims; and (c) led to adoption.

**Table 4.** Univariable logistic regression models for association between project outcome of adoption (dependent variable) and contextual, input, and process factors (independent variables).

|  | n<sub>1</sub> | n<sub>2</sub> | Odds Ratio | 95% CI |
| --- | --- | --- | --- | --- |
| **Contextual factors** |  |  |  |  |
| Inpatient | 22 | 3 | 0.5 | 0.1 – 2.3 |
| First QI project + | 27 | 2 | 0.2 | >0.1 – 0.9 |
| Time officially sanctioned + | 20 | 7 | 5.2 | 1.2 – 23.4 |
| Funding required | 10 | 3 | 2.1 | 0.4 – 10.4 |
| Funding available | 5 | 0 | # | # |
| Made budget plan | 5 | 1 | 1.1 | 0.1 – 10.6 |
| **Input factors** |  |  |  |  |
| Project lead: non-clinical staff | 8 | 1 | 0.6 | 0.1 – 5.1 |
| Project lead: non-managerial staff | 18 | 3 | 0.8 | 0.2 – 3.4 |
| Team size | - | - | 1.4 | 0.7 – 2.7 |
| Staff turnover | 22 | 4 | 0.9 | 0.2 – 3.6 |
| **Process factors** |  |  |  |  |
| Engaged clinical team lead | 28 | 6 | 1.4 | 0.3 – 5.5 |
| Engaged stakeholders (staff not in project team) | 14 | 5 | 3.7 | 0.9 – 15.5 |
| Engaged service users ++ | 10 | 5 | 7.4 | 1.6 – 34.9 |
| No. of outcome measures + | - | - | 3.2 | 1.1 – 9.8 |
| No. of primary drivers + | - | - | 2.7 | 1.3 – 5.9 |
| No. of secondary drivers + | - | - | 1.5 | 1.1 – 1.9 |
| Aims quantified | 35 | 10 | # | # |
| Primary drivers tagged with measures ++ | 13 | 6 | 7.5 | 1.7 – 33.7 |
| Secondary drivers tagged with measures + | 11 | 5 | 6.0 | 1.3 – 27.2 |
| Balancing measures ++ | 10 | 5 | 7.4 | 1.6 – 34.9 |
| No. of PDSA completed + | - | - | 1.5 | 1.1 – 2.2 |
| PDSA that completed >1 cycle ++ | 13 | 6 | 7.5 | 1.7 – 33.7 |
| PDSA with documentation ++ | 13 | 9 | 85.5 | 8.5 – 860.2 |
| Data collected before implementing change idea ++ | 9 | 5 | 9.5 | 1.9 – 47.6 |
| Median value of random variation in outcome measures + | 12 | 5 | 5.0 | 1.1 – 22.0 |
| Median value of random variation in process measures | 4 | 4 | # | # |
| Median value of random variation in balancing measures | 2 | 2 | # | # |
| Data collected after implementing change idea ++ | 13 | 6 | 7.5 | 1.7 – 33.7 |

n<sub>1</sub>: total number of projects that satisfy the condition described by the independent variable
n<sub>2</sub>: total number of projects in n<sub>1</sub> that led to a change in routine practice (adoption).

The odds of success were lower for projects in inpatient settings and those that were first attempts of the team undertaking them. However, only the latter showed a statistically significant impact with a large effect size (OR = 0.2, i.e. odds were five times lower). Project funding (need / availability / planning) appeared to have little impact, but there were too few funded projects for stable estimates. The odds of success were five times higher when time was officially sanctioned for the project (OR = 5.2). This suggests that whilst the initial years of QI applications presented major challenges, this might be mitigated by time commitments that were officially sanctioned. In terms of QI governance, advancing the organisational culture of QI practice is likely to require formal commitment of resources like staff time for training and project development.

The type of staff role in team leadership, team size, and disruption due to staff turnover did not show a reliable impact on project outcomes. Support available from the resident team of QI specialists might have mitigated the influence of these input factors. Relative to contextual and input factors, process factors showed more consistency in having an impact. Projects that engaged service users had much higher odds of success (OR = 7.4). At problem definition stage, careful application of the driver diagram methodology showed a similar impact. For instance, the odds of success increased with the number of primary drivers identified (OR = 2.7), particularly when drivers were clearly tagged with measures (OR = 6.0 – 7.0). At PDSA stages, implementation fidelity of rapid learning cycles was crucial. For instance, the odds of success were much higher for projects that completed more than one PDSA cycle (OR = 7.5), particularly when accompanied by documentation (OR = 85.5). Projects that collected data before and after implementing change ideas also had much higher odds of success (OR = 7.5. – 9.5). The processes that underpin a QI project drive the culture of continuous improvement. In terms of QI governance, a routine view of process factors can illuminate on organisation-wide training priorities and strategies.

## DISCUSSION

In this study, we explored what could be routinely reported to the executive board to give an organisation-wide picture that offers accountability and feedback for advancing QI in practice across diverse specialist care services.

In terms of project outcomes, we recommend tracking whether projects led to a change in routine practice (i.e. adoption). As a fundamental goal of QI (Batalden and Davidoff, 2007), adoption rates offer a conceptually sound summary across heterogenous projects. They can be compared between organisational units and monitored over time. Our retrospective data also underscores the value of tracking basic outcomes such as project completion and whether project aims have been achieved. Comparing the rates of these outcomes and adoption can inform on the extent to which QI is making inroads across the organisation.

We found that overall project duration and time invested in meetings and correspondence are data with minimal information retrieval burden. Across a diverse range of projects, time invested by staff is an opportunity cost that could serve as a common denominator for return on investment. In contrast, we encountered major challenges in measuring benefits. For instance, not every project captured cost savings because it is often not an immediate focus of QI in mental health services (which was the context of the NHS Trust of this study). On the other hand, while all projects arguably contributed to capacity building in a culture of continuous improvement, these broader benefits were poorly captured retrospectively. Routine monitoring is needed to bring about systematic data accrual. By increasing the visibility of growth in a culture of continuous improvement, the executive board can be informed by a broader focus on the value of QI beyond cost savings.

With data on what might have an impact on project outcomes, our findings demonstrated how feedback measures can help steer the focus when accountability measures are reported. This is crucial because accountability data tends to be used for performance management and thus is rich in potential for blame (Armstrong et al., 2018). By complementing accountability reports, feedback measures can shift away from blame allocation by anchoring focus on learning and resource allocation. For instance, routine monitoring of whether time commitments for QI projects were officially sanctioned can flag a need for the executive board to shape the organisational context for supporting staff and project development. Project success rates are also likely to benefit from organisational mandate for service user involvement in QI. Consistent with the wider literature on QI in mental health (Robert et al., 2003), our data showed that service user involvement has a large impact on project outcomes. However, different models of involvement exists (Armstrong et al., 2013). Thus, there is value for more granular data to identify optimal engagement strategies.

As structured interventional experiments for testing changes, iterative learning cycles in PDSA help translate ideas into action (Reed and Card, 2016). Improvement work is less likely to succeed if iterative cycles are too few (Ogrinc and Shojania, 2014). Furthermore, a transparent, data-driven approach is paramount (Birdas et al., 2019), otherwise project teams may get stuck in the ‘Do’ phase of a PDSA, or reach no actionable insight in the ‘Study’ phase. QI training should not over-emphasise the conceptual simplicity of PDSA (Reed and Card, 2016), because it involves new ways of working and achieving high fidelity is a gradual and negotiated process (McNicholas et al., 2019). This learning process needs to be driven by psychological safety (Carmeli et al., 2009). Engaging the executive board with a routine view of PDSA fidelity may thus help galvanise leadership efforts essential for fostering an organisational climate of psychological safety and enhancing engagement with QI (Nembhard and Edmondson, 2006).

### Study limitations

This study set out to explore an organisation-wide picture of QI projects and their impact. The study was carried out within a single UK mental health Trust, which may limit its generalizability to other healthcare systems and physical care providers. While we defined adoption as bringing about a change in routine practice, the process of retrospective information retrieval posed major challenges in obtaining a wider range of indicators to illuminate the impact. Furthermore, the absence of an established definition of ‘success’ of in QI (Morganti et al., 2012) meant that the resident QI team had to rely on their own judgement when selecting a purposive sample. Coupled with retrospective recall difficulties, sampling bias is likely to have an impact on study findings. Notably however, our survey-based findings resonate with those from in-depth qualitative studies in terms of the process factors that matter for project outcomes (Woodcock et al., 2019, Taylor et al., 2014, Reed and Card, 2016, McNicholas et al., 2019). This suggests that the skilful application of QI can be routinely monitored as an indication of cultural transformation and QI success.

While several process factors showed an apparent impact on project outcomes, contextual and input factors generally showed little impact. This is likely to be due to limitations in the scope of our survey rather than evidence that these latter aspects are not important. It is well-established in improvement science literature that contextual factors play a prominent role (Kaplan et al., 2012). However, the sheer number of variables and the unpredictability of their interactions make it hard to predict the distal impact of contextual factors on project outcomes (Braithwaite, 2018). Consequently, the need to minimise data accrual burden led to a narrow scope of contextual and input factors. Instead, we prioritised the scope of process factors (e.g. fidelity of data and measurement plans), to illuminate proximal influences that are potentially amenable to staff training and interventions.

## CONCLUSION

To advance with a long-term perspective of QI success and cultural transformation, healthcare organisation executive boards need an organisation-wide overview that can be routinely made visible with minimal technical expertise. Setting up a routine monitoring framework can offer more timely vigilance than designing an elaborate retrospective evaluation, which some argued, is akin to attempting to “drive by looking in the rear-view mirror” (Hamblin and Shuker, 2020). Practically, routine monitoring can also improve data quality by alleviating information retrieval difficulties and ensuring systematic data accrual. Operationally, respondent burden can be alleviated by spreading data collection over multiple occasions. Short and well-timed surveys can optimise relevance and thus the acceptability of data collection. Data can also be collected from different types of respondents (e.g., project team lead, sponsor, resident QI team). Well-targeted respondents will provide the most accurate data if only the most relevant questions are asked, thereby also minimising respondent burden.

Designing a routine reporting framework is an iterative process involving continual dialogue with frontline staff and improvement specialists to navigate data accrual demands (Gardner et al., 2018). Across diverse project aims, there is a need to identify themes that hold core relevance in QI. For an executive board, this framework should generate strategic insights that can inform resource commitments in QI. For staff and improvement specialists, routine reporting should offer a feedback loop to support engagement and learning, finding the right focus (Soong et al., 2020) and applying QI with fidelity (Knudsen et al., 2019, Reed and Card, 2016). Developing routine reporting can be an asset for improving practice (Dixon-Woods, 2019), and honing the implementation science of QI in healthcare.

## Data Availability

Data are available upon request from the corresponding author

