## Supplementary material for "Evaluating quality improvement at scale: routine reporting for executive board governance in a UK National Health Service organisation": online supplement

* 1. QI Lead

- [redacted]
- [redacted]
- [redacted]
- [redacted]
- [redacted]
- [redacted]
- [redacted]

* 2. Borough

- Croydon
- Southwark
- Lambeth
- Wandsworth
- Lewisham

* 3. Service

- Community
- In-patient

* 4. CAG

- CAMHS
- Psychosis
- MHOAD
- Other (please specify)

* 5. Year of project

2015 2018

* 6. Month of project

1 12

* 7. Project life span (approx. in weeks)

0 — 52

* 8. Project status

- Closed
- Terminated at S
- Terminated at P
- Terminated at A
- Terminated at D

* 9. Was the project affected by staff turnover?

- No
- Yes

* 10. Aim achieved?

- Yes
- No

* 11. After the aim was achieved, how many weeks was it sustained before the project is closed?

0 — 12

* 12. After project completion, were the change ideas adopted in practice?

- Yes
- Not known
- No

* 13. Did this project trigger similar projects at other sites?

- Yes
- Not known
- No

* 14. Estimated no. of meetings with SLaM QI

0 — 10 — 20

* 15. Estimated no. of email/phone contact with/from SLaM QI

0 50 100

* 16. Estimated total hours of commute incurred by SLaM QI

0 10 20

* 17. Estimated total travel expenses incurred by SLaM QI

0 20 40

* 18. Estimated total hours of staff time officially sanctioned for QI project

0 20 40

* 19. Estimated total hours of staff time taken

0 20 40

* 20. Was funding available for the QI project?

- Yes
- No

* 21. Was funding required for the QI project?

- Yes
- No

* 22. Did the project team make a budget plan?

- No
- Yes

* 23. Is it possible to quantify the improvement in terms of cost savings?

- No
- Yes, but not attempted
- Yes

* 24. Role of the team leader in this QI project?

- They were directly involved.
- Approached, but no apparent input.
- Not directly involved, but consulted.
- Not known.

* 25. Role of key stakeholders (not in project team)?

- They were directly involved.
- Approached, but no apparent input.
- Not directly involved, but consulted.
- Not known.

* 26. Role of service users

- They were directly involved (not as beneficiaries).
- Approached, but no apparent input.
- Not directly involved, but consulted.
- Not known.

* 27. Is this the first QI project developed by this site/service?

- Yes
- Not Known
- No

* 28. How many staff members were there in this QI project team?

0 5

* 29. How many project team members received Foundation QI training?

0 5

* 30. Band of employment of the project lead.

1 9

* 31. Professional role of project lead

- Professional Services
- Clinical
- Non-clinical

* 32. Highest band of employment in this project team?

0 9

* 33. Lowest band of employment in this project team?

0 9

* 34. How many more QI projects were developed by any of these staff members since?

0 2 4

* 35. Did the project team disseminate the results?

- [ ] At the local site/service
- [ ] Not Known
- [ ] Beyond the local site/service

* 36. Did the project team disseminate the results?

- [ ] In a form of publication (e.g newsletter)
- [ ] Not known
- [ ] At a conference

* 37. Estimated no. of service users who directly benefited from the change introduced.

0 20 40

* 38. Estimated no. of staff who directly benefited from the change introduced.

0 20 40

* 39. The project aligns with Trust Quality priorities for

- [ ] Patient Safety: Reduce the use of restrictive interventions applied to service users.
- [ ] Patient Safety: Safer staffing
- [ ] Patient Safety: Risk Assessments
- [ ] Clinical Effectiveness: Physical healthcare screening
- [ ] Clinical Effectiveness: Care planning
- [ ] Clinical Effectiveness: Developing electronic systems to improve the delivery of care
- [ ] Patient Experience: Reducing the number of Acute out of area treatments
- [ ] Patient Experience: Carer's assessments and associated care plan
- [ ] Patient Experience: Quality of environments and food within in-patient services

* 40. No. of outcome measures attached to the Aim identified in the driver diagram.

0 2 4

* 41. Were the target outcomes quantified in the Aim?

- ( ) No
- ( ) Yes

* 42. How many primary drivers were there in the driver diagram?

0 5 10

* 43. No. of primary drivers with a measure attached to it

0 5 10

* 44. How many secondary drivers were there in the driver diagram?

0 5 10

* 45. No. of secondary drivers with a measure attached to it

0 5 10

* 46. No. of change ideas identified in the driver diagram

0 5 10

* 47. No. of change ideas tested and retained

0 5 10

* 48. No of change ideas tested and discarded

0 5 10

* 49. No. of PDSA constructed for the entire project

0 5 10

* 50. No. of PDSA with documentation to enable others to replicate what was done

0 5 10

* 51. No. of balancing measures introduced to check for unintended consequences

0 5 10

* 52. No. of PDSA that completed 1 cycle (i.e. concluded with an informed action)

0 5 10

* 53. No. of PDSA that completed more than 1 cycle

0 5 10

* 54. Was there clear evidence to suggest that an improvement is needed?

- No
- Yes

* 55. How many times was data collected before implementing change ideas?

0 5

* 56. No. of weeks data was collected before implementing change ideas?

0 5 10

* 57. Was the median value of random variation established for outcomes?

- No
- Yes

* 58. Was the median value of random variation established for process measures?

- No
- Yes

* 59. Was the median value of random variation established for balancing measures?

- No
- Yes

* 60. How many times was data collected after implementing change ideas?

0 5

* 61. No. of weeks data was collected after implementing change ideas.

0 10
